# Meta-evaluation of a whole systems programme, ActEarly: A study protocol

**DOI:** 10.1101/2023.01.09.23284353

**Authors:** Liina Mansukoski, Bridget Lockyer, Amy Creaser, Jessica Sheringham, Laura Sheard, Philip Garnett, Tiffany Yang, Richard Cookson, Alexandra Albert, Shahid Islam, Robert Shore, Aiysha Khan, Simon Twite, Tania Dawson, Halima Iqbal, Ieva Skarda, Aase Villadsen, Miqdad Asaria, Jane West, Trevor Sheldon, John Wright, Maria Bryant

## Abstract

**Introduction:** Living in an area with high levels of child poverty predisposes children to poorer mental and physical health. ActEarly is a 5-year research programme that comprises a large number of interventions (>20) with citizen science and co-production embedded. It aims to improve the health and well-being of children and families living in two areas of the UK with high levels of deprivation; Bradford in West Yorkshire, and the London Borough of Tower Hamlets. This protocol outlines the meta-evaluation (an evaluation of evaluations) of the ActEarly programme from a systems perspective, where individual interventions are viewed as events in the wider policy system across the two geographical areas. It includes investigating the programme’s impact on early life health and well-being outcomes, interdisciplinary prevention research collaboration and capacity building, and local and national decision making.

**Methods:** The ActEarly meta-evaluation will follow and adapt the five iterative stages of the ‘Evaluation of Programmes in Complex Adaptive Systems’ (ENCOMPASS) framework for evaluation of public health programmes in complex adaptive systems. Theory-based and mixed-methods approaches will be used to investigate the fidelity of the ActEarly research programme, and whether, why and how ActEarly contributes to changes in the policy system, and whether alternative explanations can be ruled out. Ripple effects and systems mapping will be used to explore the relationships between interventions and their outcomes, and the degree to which the ActEarly programme encouraged interdisciplinary and prevention research collaboration as intended. A computer simulation model (“LifeSim”) will also be used to evaluate the scale of the potential long-term benefits of cross-sectoral action to tackle the financial, educational and health disadvantages faced by children in Bradford and Tower Hamlets. Together, these approaches will be used to evaluate ActEarly’s dynamic programme outputs at different system levels and measure the programme’s system changes on early life health and well-being.

**Discussion:** This meta-evaluation protocol presents our plans for using and adapting the ENCOMPASS-framework to evaluate the system-wide impact of the early life health and well-being programme, ActEarly. Due to the collaborative and non-linear nature of the work, we reserve the option to change and query some of our evaluation choices based on the feedback we receive from stakeholders to ensure that our evaluation remains relevant and fit for purpose.

## Introduction

Living in an area with high levels of child poverty predisposes children to poorer mental and physical health outcomes due to exposures to economic, physical, built, cultural, learning, social, and service environmental risk factors [1, 2]. With health inequalities anticipated to widen over time [3], various interventions, initiatives and policies have been implemented aimed at reducing health inequality gaps (e.g. providing families with income support, changes in environmental infrastructure and public health campaigns), with varying impact [4, 5].

ActEarly is a UK Prevention Research Partnership (UKPRP) funded research consortium focused on improving the health and well-being of children and families living in two areas with high levels of deprivation; Bradford in West Yorkshire, and the London Borough of Tower Hamlets [6]. ActEarly is unique, in that it is a series of interlinked interventions, embedded with citizen science and co-production of research with local communities, across two complex systems [6]. The 5-year programme, whose implementation began in September 2019, creates testbeds of upstream interventions within whole-system city settings, supporting intervention identification, implementation and evaluation [6]. A ‘whole-system city setting’ can be thought of as a complex adaptive system consisting of multiple interconnected, emergent and dynamic parts that are open to influences both from inside and outside of the system [7]. ActEarly aims to: 1) establish a prevention research consortium that unites broad transdisciplinary expertise (e.g., economics, urban design, transport, education, housing, social justice and welfare) with the public, policy leaders and practitioners from across the two areas; 2) identify, co-produce and implement system-wide early life upstream prevention solutions; 3) provide efficient data platforms and methodological expertise enabling robust population-scale evaluation of the impact of interventions on environments, health related behaviours and interlinked health, educational, social and economic outcomes; and 4) evaluate, refine, replicate and disseminate the City Collaboratory approach as a model for addressing upstream determinants of health and inequality [6].

To date, over 40 individual projects have been launched under the ActEarly umbrella, including over 20 interventions targeting the ActEarly themes: ‘healthy places’, ‘healthy learning’, ‘healthy livelihoods’, ‘food and healthy weight’ (cross-cutting theme) and ‘play and physical activity’ (cross-cutting theme) [6], with citizen science and co-production of research with local communities across the two study sites [8, 9]., It is anticipated that the number of interventions and projects will increase as the project continues. The nature of complex systems inevitably involves a wide range of target areas which means resource has been spread relatively thinly across the child health system, and ActEarly funding did not cover the cost of interventions.

In the short term, rather than a sole focus on large and measurable improvements in health and wellbeing, our evaluation of ActEarly will explore changes across various processes of researcher collaboration, local authority partnership working, co-production, citizen science, and data linkage. This type of evaluation is crucial for understanding the mechanisms of action behind an intervention and understanding how and why an intervention is (in)effective [10]. The evaluation of the ActEarly programme is unique in that it will require a meta-evaluation (an evaluation of evaluations), given the >20 dynamic interventions included within the system [6, 11]. It necessitates a systems dynamic perspective, where individual interventions are viewed as events and wider policy system change across the two study areas. This is because, in any research programme, programme outcomes may be a result of the context, rather than the implementation of the programme alone [12]. The evaluation of complex systems interventions requires a hybrid research design, in which more than one overall approach is combined and tailored to support the evaluation [13]. This builds on realist methodology, and further acknowledges that focusing on events at a single time point cannot sufficiently describe how the intervention interacts with changes in the wider complex adaptive system [12, 14]. Instead, a two-phase approach is needed where first, the existing system is described, including hypothesising how the intervention may change it (i.e., theory of change, logic models). In the second phase, the changes resulting from the intervention and their interaction with the wider system is described [14].

The meta-evaluation of ActEarly itself is complex, because the projects that are part of the research portfolio encompass a wide range of different types of interventions, with a range of stakeholders, working across different areas (including the transport system, educational system, health system and food system). Defining which systems ActEarly targets is therefore challenging, and as such, it has been more useful to think of the impact of ActEarly as something that targets the wider child health policy system of the geographical areas, with individual subsystems (e.g., healthcare) as interlinked parts. It could be argued that the target of ActEarly is a type of ‘meta-system’ - the wider policy environment in which decisions are made to direct resources to different and often competing causes. ActEarly therefore contributes to the *environment*, or context, in which other systems exist. The implication of this for the design and focus of the meta-evaluation is that the methodology of the evaluation needs to have sufficient flexibility to capture wide ranging and even unexpected changes in the system. Many of these may not be evident in the study design phase and may only be discovered during the evaluation process. This is because in a complex system, it is difficult to foresee the outcome of different elements interacting (interventions, stakeholders, wider political and policy environment).

The difficulty of evaluating complex adaptive systems is acknowledged, and as a result the ‘Evaluation of Programmes in Complex Adaptive Systems’ (ENCOMPASS) framework has previously been developed [7]. The ENCOMPASS framework outlines five iterative stages that can be applied to complex public health programmes, such as the ActEarly programme (e.g., from defining system boundaries to measuring programme outcomes) [7]. Consequently, the aim of this protocol is to outline how the meta-evaluation of the ActEarly research programme will be conducted, by using and adapting the ENCOMPASS framework to meet the aims and scope of ActEarly.

## Methods

### The ENCOMPASS framework

The ActEarly meta-evaluation will follow and adapt the ENCOMPASS-framework [7], that has been developed to guide the evaluation of public health programmes in complex adaptive systems. The ENCOMPASS framework consists of five stages: 1) adopting a system dynamics perspective on the overall evaluation design; 2) defining the system boundaries; 3) understanding the pre-existing system to inform system changes; 4) monitoring dynamic programme output at different system levels; and 5) measuring programme outcome and impact in terms of system changes [7]. How the ENCOMPASS framework will inform the meta-evaluation of the ActEarly programme is described below, but it is important to note that the process of evaluation using this framework is non-linear, that is, stages may be revisited as the process evolves.

### Stage 1 - Adopting a system dynamics perspective on the overall evaluation design

Stage 1 of the ENCOMPASS-framework involves (a) specifying a logic model, (b) specifying the stages and aims of the evaluation, (c) framing evaluation questions, and (d) forming an evaluation team. The evaluation team has already been formed based on the individual’s expertise and knowledge, therefore stages 1a-c will be discussed in this section.

#### Stage 1a. Specifying a logic model

The programme theory of ActEarly was initially developed in the consortium building phase in 2018 and the logic model highlights that, through ActEarly activities (research, data initiatives, citizen science and community engagement) knowledge and evidence will be gathered and lead to increased awareness and perceived relevance in early years health and well-being at the local and national level. Successful initiatives and changes in decision making are proposed to eventually result in reductions in inequalities and non-communicable disease (Fig 1). The idea underpinning the model is that changing any one element within the system is not sufficient to see the desired changes in outcomes, but by enacting systems wide change, the cumulative effect of the programme will lead to measurable impact.

**Fig 1.**
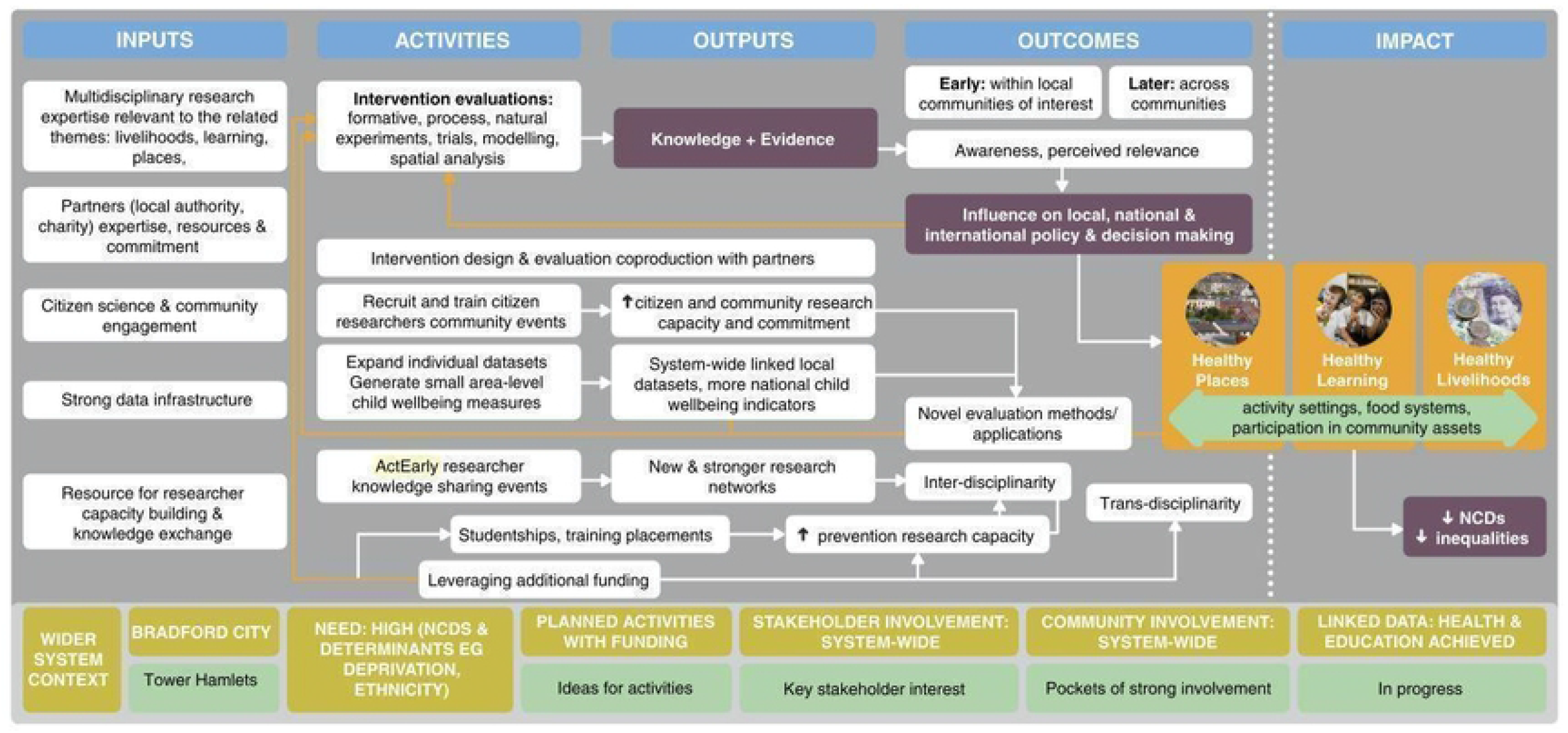
ActEarly original logic model

The initial logic model was developed in 2019 (Fig 1), and revised in 2022 (Fig 2) to highlight the pathways to outcomes and impact the research programme aims to achieve, following a period of implementation and reflection. The consortium has produced a description of the logic model in an audio-visual format [15]. The inputs in the model reflect what was already in place, whereas the activities are what we envisioned ActEarly would contribute to within the system. The two study sites are shown to be similar in terms of some of the wider system context, but start off at different stages in terms of readiness for ActEarly (e.g., there were more planned activities and system wide stakeholder engagement in place in Bradford at the start of the project). Interventions and individual evaluations are intended to increase knowledge, evidence, and awareness and perceived relevance of early life health and well-being, first within the local communities and later across communities. As the project matures, we expect that this may lead to increased influence on policy making. It is anticipated that important feedback loops will exist alongside this main pathway to impact, and that the other ActEarly activities of co-production, citizen science and developing strong linked datasets across both sites will contribute to the impact that the project has.

**Fig 2.**
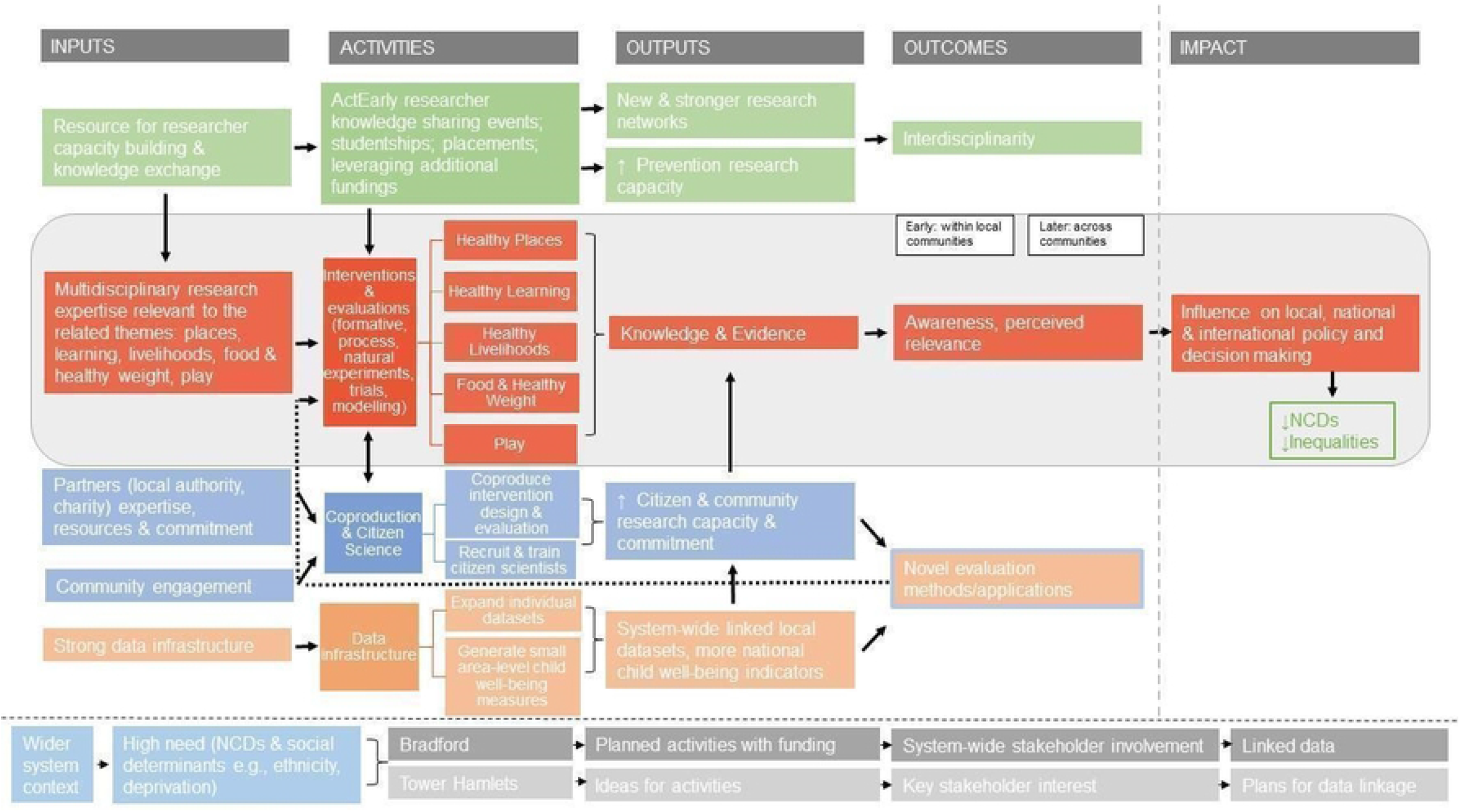
ActEarly revised logic model

The revised logic model was used to develop the questions the meta-evaluation seeks to answer. The core members of the evaluation theme of ActEarly held two online workshops to discuss the model and potential questions, and then sought feedback from the wider team before agreeing on the final set of questions (see Stages 1b and 1c).

#### Stage 1b. Specifying the stages and aims of the evaluation

The five iterative stages outlined by the ENCOMPASS framework will be used and adapted to define the ActEarly programme’s system boundaries, understand the pre-existing and current system ActEarly exists within to inform system changes, monitor the dynamic programme outputs of ActEarly at different system levels and measure ActEarly’s outcomes and impact on system changes.

#### Stage 1c. Framing evaluation questions

The evaluation questions were based on the intended outputs, outcomes and impacts outlined in the updated ActEarly logic model (Figure 2). The questions were developed to capture if and how these are realised and identify interacting contextual factors. The questions were queried and refined by the ActEarly Evaluation team over the course of several meetings. Drawing on a systems perspective, the evaluation questions that this meta-evaluation will address are:

1. What major external contextual factors were likely to influence the implementation of ActEarly?
2. How, and to what extent, was ActEarly implemented as intended?
  a. What effect has ActEarly had on interdisciplinary and prevention research collaboration?
  b. What effect has ActEarly had on interdisciplinary and prevention research capacity?
  c. To what extent has ActEarly improved citizen science and community research capacity and commitment across both study sites (e.g., through extended networks)?
  d. To what extent has ActEarly contributed to system-wide linked local datasets?
  e. Did ActEarly influence local and national decision making and if so, how?
  f. To what extent has the evidence from ActEarly intervention evaluations been acted on? (e.g. investment, disinvestment, continuation)
  g. Was ActEarly able to intigate interventions that will persist beyond the life of the project? If yes, how? If not, why?
3. Is there any evidence that ActEarly has started to have a meaningful impact on early life health and well-being outcomes?
  a. Have ActEarly intervention evaluations reported any measurable changes in the outcomes included in the public health core outcome sets for early years (COS-EY)?
  b. Are there likely to be any long-term changes in outcomes due to ActEarly activities?

### Stage 2 - Defining the system boundaries

The second stage of the ENCOMPASS-framework involves defining the system boundaries to determine what lies ‘inside’ and ‘outside’ of the system. This process guides what is and what is not included in the scope of the meta-evaluation based on the programme’s purpose and determining who and what is part of the system. To address this, it is important to describe the environment surrounding the evaluation.

#### Environment surrounding the evaluation

ActEarly takes place within a complex public health policy environment across the two local authority areas of the Bradford District and the Borough of Tower Hamlets in London. While the two areas share similarities in terms of certain population demographics (area level deprivation, health outcomes, ethnic diversity), we describe each site separately and in detail below.

##### Bradford

The City of Bradford Metropolitan District Council (CBMDC) is the 5th largest metropolitan District in England, with over 540,000 residents [16]. The district is known for its young and diverse population, with 30% of the population aged 19-years or younger and 36% of people from ethnic minorities and over 150 languages spoken [16]. Notably, there is a large population of Pakistani origin in Bradford (20%) [16]. Within the district, there are considerable health inequalities between affluent and deprived areas [17]. Bradford as a district continues to face major challenges and has one of the lowest healthy life expectancies in England, which is predominantly lower than the national average for males and females, particularly in more deprived areas [17]. Half of Bradford residents live in council wards where unemployment is above average [18], and child educational attainment is below regional and national levels, particularly for children who are deemed “persistently disadvantaged (on free school meals for >80% of their school life) [19]. Child poverty is high with 30% of children in absolute low income families (living in households with an income below 60% of median income in some base year) compared to the England average of 15.3% [17], and families living in poor housing conditions [20].

The district’s local authority, the CBMDC, is a crucial actor in setting the priorities of the early years public health policy system by managing and planning built environments, access to education and skills, as well as providing transport, sport and leisure, public health, welfare, and other support services. Priorities and principles that guide the resource allocation between these areas follow in part from the changes in the priorities of locally elected democratic leaders. Currently, the district’s plans in terms of health and early years policy priorities are set out in the Bradford Council Plan 2021-2025 [21]. Key principles detailed within this plan include embedding prevention and early help across the system, with one key priority being ‘Better Health, Better Lives’ which aims to improve the health and wellbeing for everyone in the district through e.g., the Living Well initiative [21].

Supporting the work of the CBMDC is the Bradford Institute for Health Research (BIHR), which was established in 2006 to support evidence-based local decision making. The BIHR houses National Institute for Health Research (NIHR) research centres including the Yorkshire and Humber NIHR Applied Research Collaboration (ARC). With support from the NIHR, BIHR led the Local Authority Research system (LARS) project which explored what a LARS for Bradford might look like [22]. Subsequently, the NIHR Unlocking Data project has started to scope whole-system data linkage for health, education, social care, crime, and housing [23]. In addition, the BIHR houses the Born in Bradford cohort study, tracking the lives of over 30,000 Bradfordians to find out what influences the health and wellbeing of families [24]. All together these initiatives engage over 50,000 Bradford residents in research activities and have led to the ambition of establishing Bradford as a ‘City of Research’. This includes aiming to recruit the world’s biggest community of health research volunteers that through their participation in research activities can support evidence-led policy making [25]. As such, the public health policy environment in Bradford can be argued to be characterised by close links between the local authority and researchers, but as of yet, it is unknown if this continued collaboration has resulted in any positive shifts or changes in early years health and well-being outcomes for the residents of the district.

##### Tower Hamlets

The Borough of Tower Hamlets is located within London and has 310,300 residents speaking 137 languages [26]. It is the fastest growing population in England, and is home to the largest Bangladeshi population in the country (1 in 3 residents) [26]. Sixty percent of the borough live in the 30% most deprived areas in England, with 4 in 10 households living below the poverty line and 23% of households rely on housing benefits to pay their rent [27]. Similar to Bradford, Tower Hamlets faces multiple public health challenges and has the 6^th^ lowest disability free life expectancy in London [28]. At a local authority level, 32% of school children in Tower Hamlets are persistently disadvantaged, and these children are 6-months behind their peers in secondary level GCSE subjects; English and Maths [19].

The strategic plan of the Tower Hamlets local authority for 2019-2022 aimed to ensure that children and young people in the borough get the best start in life and realise their potential – this was part of fulfilling the council’s priority of “*people are aspirational, independent and have equal access to opportunity*” [29]. Following the change of political administration in May 2022, the Strategic Plan for 2022-2026 sets out amongst its priorities tackling the cost of living crisis (with particular focus on tackling poverty and food insecurity), housing and education [30]. To achieve these aims, the local authority is committed to research informed policy making and has a track record of collaborations and links with higher education partners and the voluntary sector. While the data and community research infrastructure in Tower Hamlets are not as linked up with the local authority as they are in Bradford, there has been an increased effort in recent years to facilitate this e.g., allowing researchers access to routine data to support council decision making, including developing data infrastructure that can support public and population health [30].

##### System boundaries of ActEarly

The updated logic model for ActEarly is the result of a co-design process that sought input from researchers across the consortium (see Stage 1a; Figure 2). As such, it is a natural starting point for setting the boundaries for the meta-evaluation. Based on the logic model, we have created criteria for inclusion and exclusion of projects in the meta-evaluation, using the format of a ‘decision tree’ (S1 Fig). Overall, we anticipate that the boundaries of the evaluation are bound to be somewhat fuzzy, as ActEarly targets a whole city environment, and not all interlinked parts can be identified in advance. If we find that our current approach seems to exclude key activities, we will revisit the boundary decisions proposed in this protocol (the decision tree; S1 Fig).

#### Defining the boundaries of the wider system

The wider systems that ActEarly is operating within, both in Bradford and Tower Hamlets are expansive and the boundaries are particularly hard to identify. Unlike the authors of the ENCOMPASS framework, ActEarly is targeting many parts of the child health system across Bradford and Tower Hamlets, rather than a specific part of it. Part of the challenge comes from ‘children’s health’ or the child health system not being a single clearly definable system, but instead a highly complex, evolving, system-of-systems. Each of those systems-of-systems will have varying significance to the different research projects and interventions. There will also inevitably be overlaps with numerous other systems, such as education. Boundaries between systems are therefore fuzzy and not fixed, and will be set to more of a pragmatic limit.

The child health system is therefore a wicked problem, without simple solutions or clear boundaries. Instead it is characterised by trade-offs between different objectives and what is possible, competing interests, incomplete information and so on. Children’s health is also highly political, and views will differ as to what is a suitable approach to solving different problems and domains of responsibility may in some cases be disputed.

This makes the formation of clear definition system boundaries for ActEarly, that can be applied systematically to determine what is in or out of scope, practically impossible. Instead, what is in or out of scope will be determined more through dialogues with stakeholders around how significant something is to children’s health (which will be guided by systems thinking approaches, such as systems mapping), and whether or not the target of investigation is within the two research areas of Bradford or Tower Hamlets. It is also likely that there will be a political dimension to these discussions, centred around if the target of investigation can be effectively influenced by ActEarly and its partners.

### Stage 3: Understanding the pre-existing system to inform system changes

To answer each of our evaluation questions, the ENCOMPASS framework recommends (a) mapping the pre-existing system in which the ActEarly programme exists, as well as (b) identifying the ‘levers of change’ within the systems identified (hypothesised to be the ActEarly activities, *e*.*g*., *knowledge sharing events, studentships, interventions, evaluations, coproduction & citizen science activities, and data infrastructure activities*; see Figure 2).

#### Stage 3a: Mapping the pre-existing and current system

The work to map the wider child health system in Bradford and Tower Hamlets is ongoing and has consisted of multiple local mapping exercises between 2018-2022, as well as interviews with people working in the system before ActEarly started. Thus, some of this work was conducted prior to the implementation of ActEarly in 2019 (2018), but also during the implementation of ActEarly (2019-2022). This mapping work provides a sense of relationships within ActEarly, and how they are developing through time. The changing relationships between ActEarly and the local communities are also captured through mapping ActEarly projects and their links with individuals and partner organisations. We will use Jessiman et al’s [31] systems map of the determinants of child health inequalities in England at the local level as a starting point to develop specific ‘child health’ maps for Bradford and Tower Hamlets. Following an initial ‘proof of concept’ exercise to review the existing child health map [31] within the ActEarly team and partners, we will conduct two workshops with key ActEarly stakeholders to consider where ActEarly is operating within the map. This will enable us to overlay ActEarly projects and other activities to highlight the areas that ActEarly is enacting on, whilst highlighting gaps in implementation. Other sources of information that will contribute to the pre-existing system’s map include smaller maps that individual ActEarly teams develop, ActEarly project logs and other records of activities to date. Although covering a period of multiple years, this process will create a description of the baseline system, as well as the ActEarly project itself.

#### Stage 3b: Identifying levers of change

The ActEarly approach assumes that new levers of change in the pre-existing system will be formed following the ActEarly activities: knowledge sharing events, studentships, placements, securing of additional funding, interventions, evaluations, coproduction & citizen science activities, and data infrastructure activities. We envisage, for instance, that the added benefit of ActEarly (over the effect of influential individuals working on their own outside the consortium), could be to join up individuals and information structures across the two local authority areas. In this case, the levers of change would be the act of bringing similar local authority areas together to solve issues that are the same or similar across both sites. The methodological approaches we intend to use to formally evaluate each lever (activity and output) from our logic model is described below under the fourth and fifth stages of the ENCOMPASS-framework which seeks to monitor dynamic programme output at different system levels, and to measure programme output and impact in terms of systems changes.

### Stage 4: Monitoring dynamic programme output at different system levels

Stage 4 of the ENCOMPASS framework involves capturing data that can be used to monitor the programme. The ActEarly meta-evaluation includes a concurrent triangulation study design where qualitative and quantitative approaches are employed simultaneously [32]. ActEarly’s evaluation will not only assess the reported impacts of ActEarly interventions, but seek to evaluate the formation, outputs, and impact of transdisciplinary collaborations. To achieve this, we will employ theory-based approaches to articulate if and how ActEarly is working to deliver change. This approach will further investigate whether, why or how ActEarly contributes to changes in the policy system, and whether alternative explanations can be ruled out. In addition to interviews and document analysis, we will employ ripple effects [33] and systems mapping [31] techniques to support the systematic analysis of multiple interventions. Details of the approach we will use to address each evaluation question (1, 2a-g, 3a-b) is displayed in Table 1 and discussed in further detail in the ‘Methods and analyses in depth’ section.

**Table 1.**
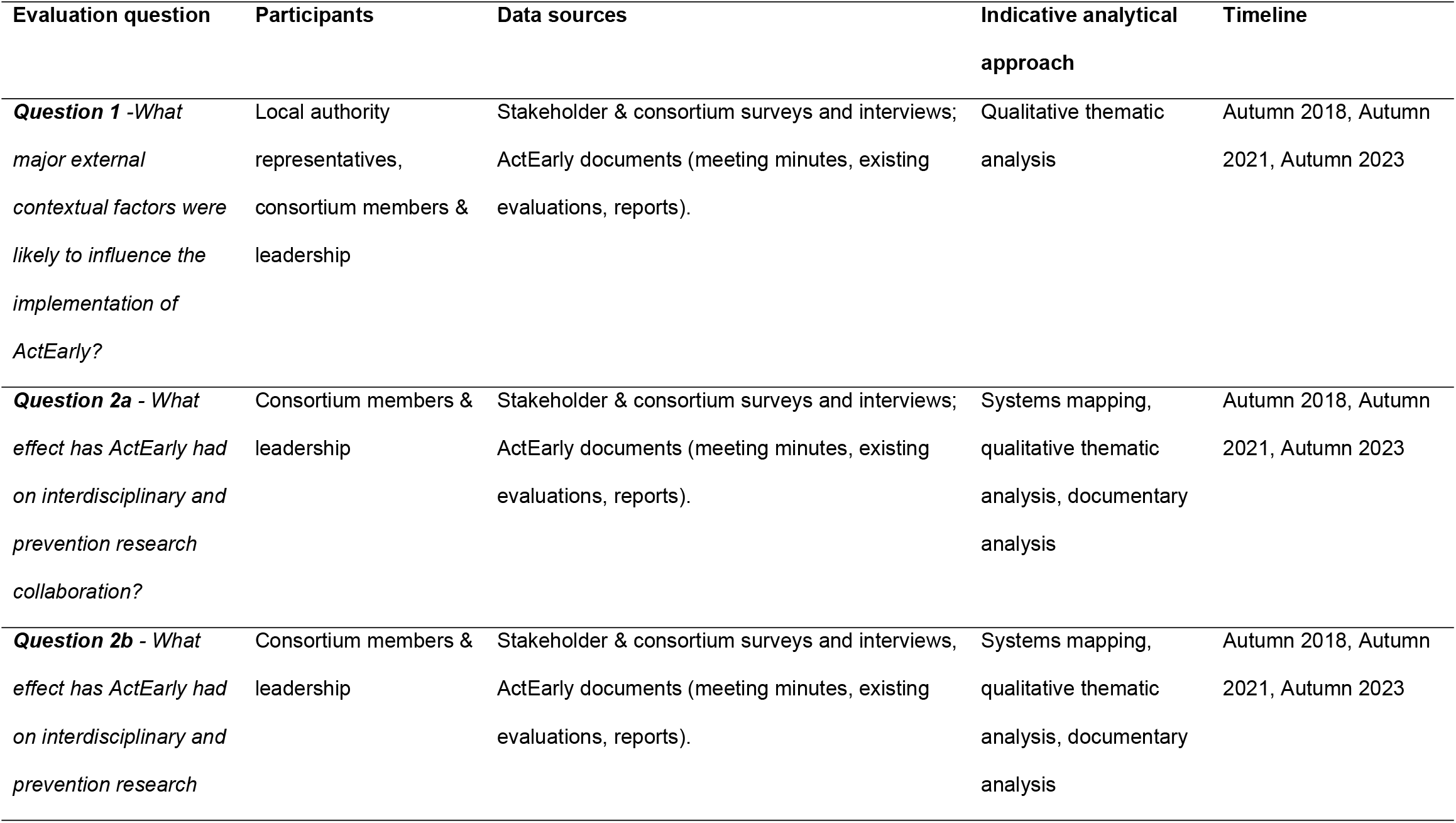
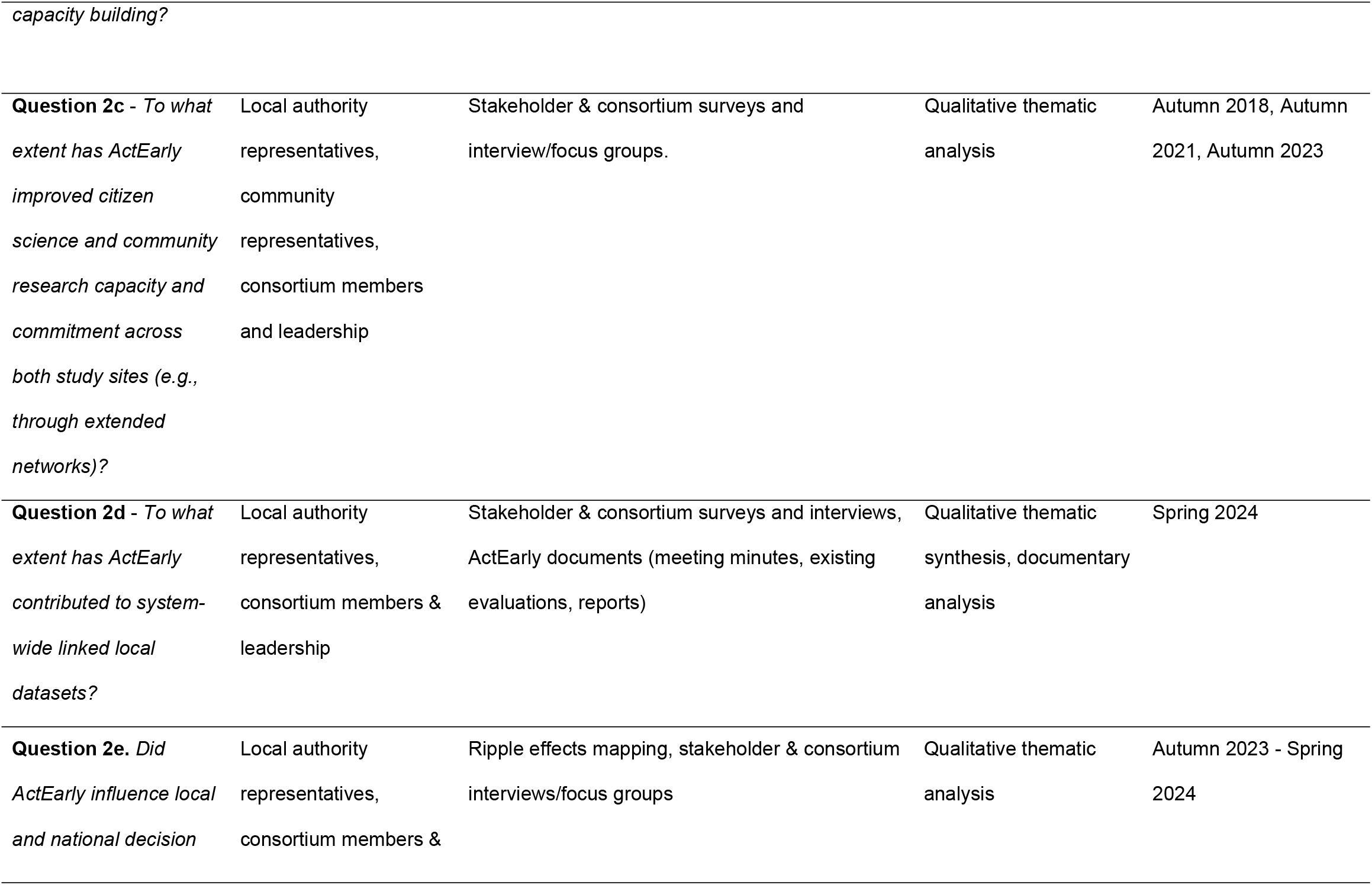
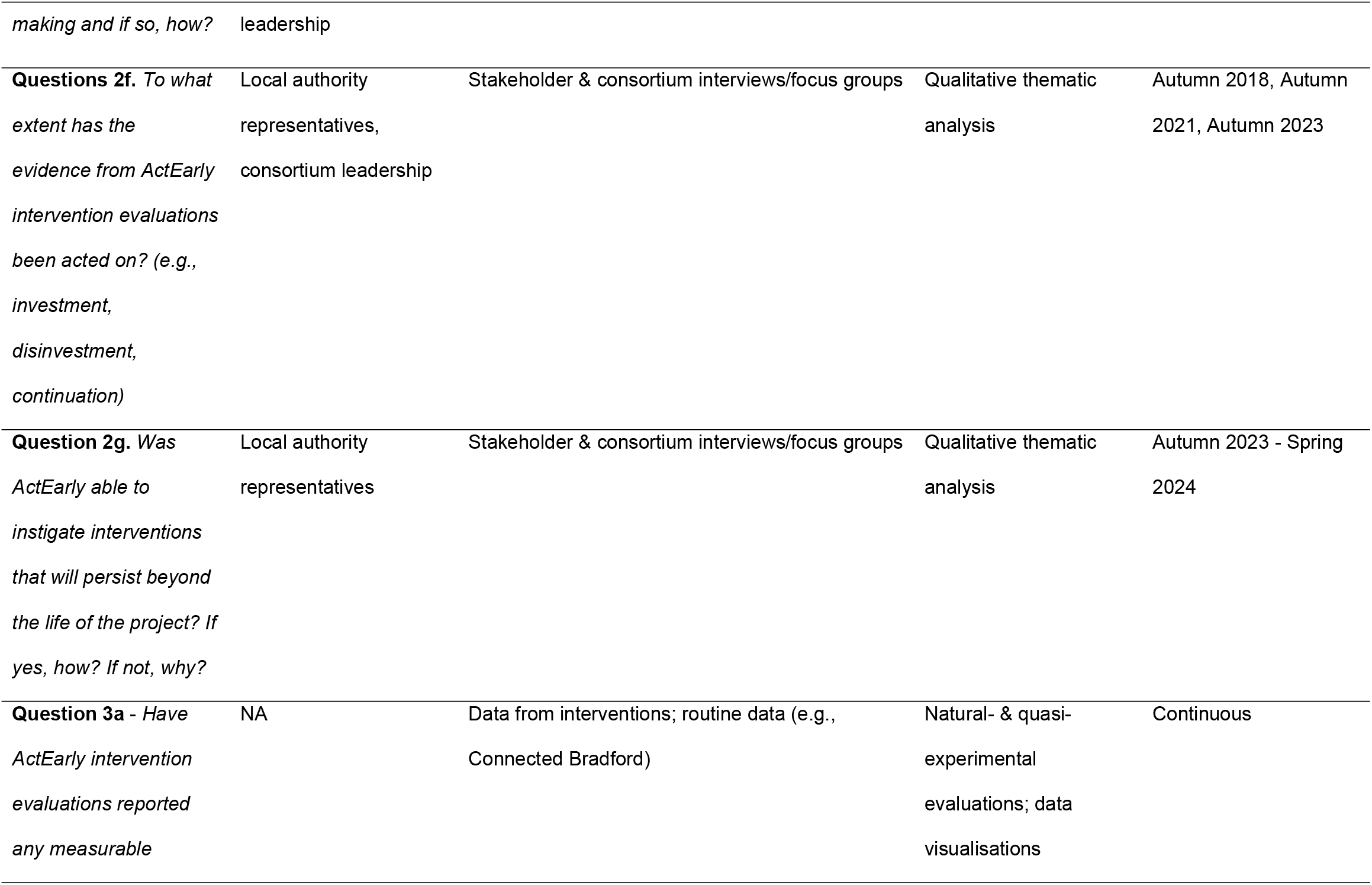
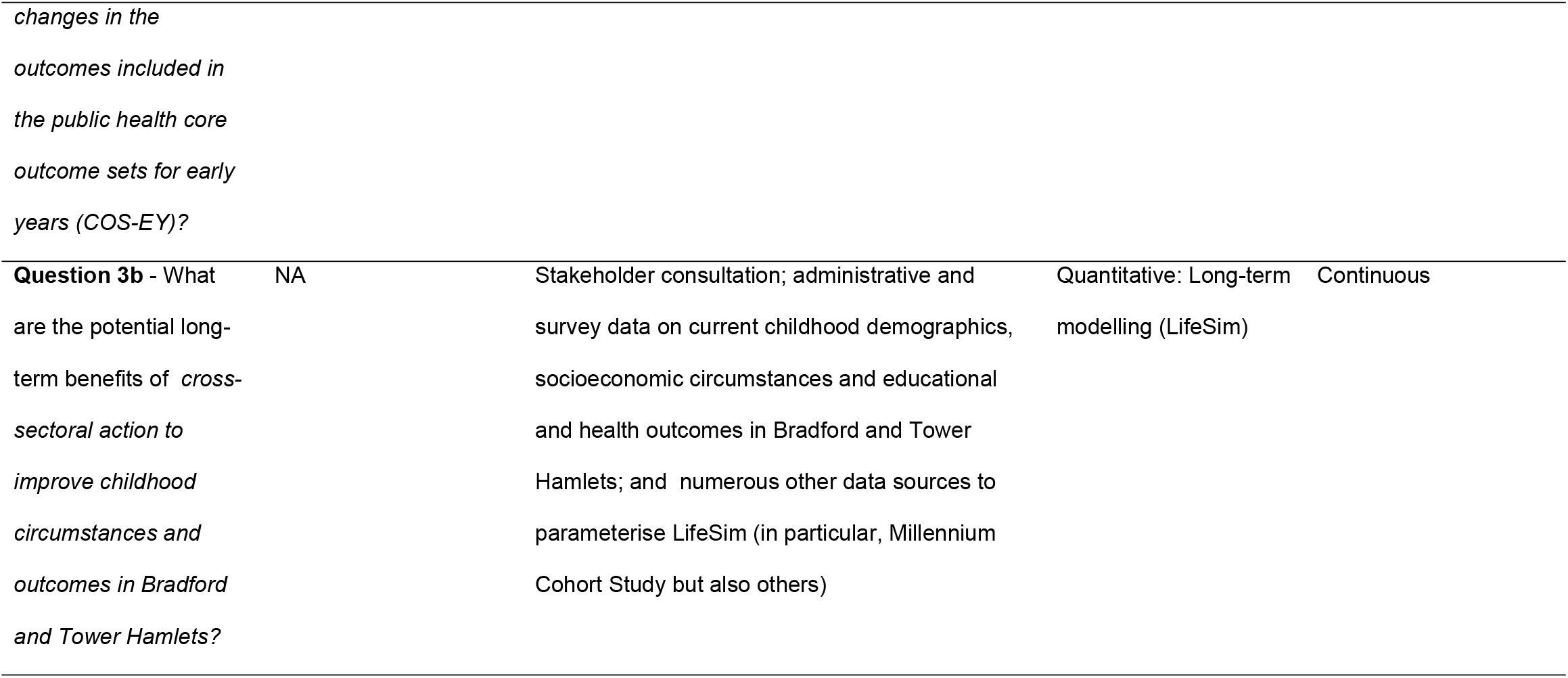
ActEarly meta-evaluation questions and their corresponding data collection and analytical approaches

Unlike conventional approaches, we will remain open to adjusting analytical approaches until closer to the end of the ActEarly project, when it is clearer what the individual projects, operating as part of ActEarly, have conducted in terms of evaluations. For evaluation of primary quantitative data (e.g. from meta-evaluation surveys), a predetermined analysis plan will be developed prior to reviewing the data.

A combination of data sources will be used to answer multiple questions (Stage 1c), including primary data that are collected for the meta-evaluation (e.g., consortium surveys and interviews), and data that are routinely collected by interventions, services or governments. Other methods will use secondary data sources from individual ActEarly projects, such as project protocols, reports, and published manuscripts. We may also utilise more unofficial source materials such as meeting notes. For question 2a, routine data collected across the two study sites will enable us to visualise patterns in key outcomes.

#### Methods and analyses in depth

##### Qualitative methods

Qualitative methods will be used to address eight of the meta-evaluation questions (1, 2a-g), either in part or exclusively. This is because one of our central aims is to understand the context of the system in which ActEarly interventions take place and the context specific impact of those interventions on the system. Thus, qualitative methods are most suitable for this exploratory and interpretative work and will use the following approaches.

#### Interviews and Focus Groups

Interviews and focus groups will be conducted in Autumn 2023 with stakeholders including local authority and consortium members as well as community members within Bradford and Tower Hamlets (*n*= approx 40). Most participants will be purposively sampled through our existing systems network mapping, which identifies individuals most connected within ActEarly. We will also purposely sample those who are less connected with ActEarly to understand the limits of ActEarly’s reach and why some members of the consortia are less involved. This work has already begun in part through the ‘Research on Research’ element of ActEarly, where interviews have been conducted with consortium members at two time points, before the project began in 2018 (*n*= 15) and at the mid-way point in 2021 (*n*= 20).

The last Research on Research data collection point will be combined with the wider meta-evaluation interview/focus group study and conducted in Autumn 2023 (*n of researchers*= approx 20). The co-production team will identify members of the community who have been involved with ActEarly interventions and an appropriate sample will be invited to participate in an interview or focus group. Longitudinal interview and focus group data will be analysed inductively through thematic analysis [34].

#### Documentary Analysis and Meeting Observations

We will conduct documentary analysis of selected ActEarly board and executive meetings and publicly available local authority meeting minutes (*n*= approx 20) between 2018-2024, in addition to capturing meetings through observations and reflective notes. We will also analyse the ActEarly annual reports (*n*= 4). Documents and notes will be compiled and analysed thematically. The findings will be triangulated with data from the interviews and focus groups in order to validate, refute, elucidate, or expand on findings across other data sources.

#### Ripple Effects Mapping

Ripple Effects Mapping (REM) can capture the wider impacts, and adaptive nature, of a systems approach [33]. It is a participatory method and will involve consortia members and other stakeholders in data gathering workshops. REM was chosen as a method because it is concerned with understanding contribution rather than attribution, and we are seeking to explore how a set of ActEarly interventions contributes towards changing an outcome or a system [33]. Approximately 2-5 workshops will be conducted towards the end of the ActEarly programme (2023/24). The aim of these workshops will be to create a visual output of ActEarly activities and interventions along a timeline, helping us to understand unintended and interlinking impacts of interventions [33]. Each workshop will be made up of around 20 participants, purposely sampled using the systems mapping network. We will use an adapted version of REM proposed by Nobles et al. [33] as a guide, and analyse the workshop’s materials through a content analysis.

##### Quantitative Methods

###### Systems mapping

A number of system mapping activities have been undertaken with more planned. Relatively detailed (at the level of interactions between individuals and organisations etc.) maps of the ActEarly project itself have been developed to understand the relationships between parts of ActEarly, and how these relationships have developed through time. This includes relationships, such as connections between people and organisations, across the different themes. Ongoing work is also being done to map the relationships between ActEarly and other child health focused projects in the two areas. These maps also include links to other organisations, such as the third sector and local government.

As part of the qualitative surveying of ActEarly members we have also produced a project social network built from an individual’s self-reported interactions with fellow project members over time, an exercise that will be repeated again during the project (see Table 1 for timelines).

These project system maps will be analysed in conjunction with maps developed from larger children’s health systems to develop a higher-level view of where ActEarly is interacting within the children’s health system at the two research sites, Bradford, and Tower Hamlets. We envision that these maps will allow us to develop a consensus with regards to boundaries and domains, that is derived from the existing literature, resulting in a meta-system map of children’s health that provides information (locally) about how ActEarly interventions interact with children’s health more broadly. This work is underway and will be reported on in due course elsewhere. Once completed network analysis methods will be applied to the systems maps as part of our mixed methods approach. Such as, community detection and core-periphery analysis to look for important sub-nets or produce clusters of areas with increased network density, and centrality measures to understand the relative importance of nodes.

###### Natural- & quasi-experimental evaluations

We will gather and synthesise information and data from individual ActEarly projects, including those with planned randomised controlled designs (e.g. JU:MP; [35]), quasi-experimental evaluations (e.g.BiB Breathes; [36]) and those with ‘pre-post’ evaluations to respond to question 3a. Quasi-experimental designs are utilised when it is not possible to randomise individuals/groups due to ethical, political, or logistical constraints [37]. Taken with a rigorous approach, quasi-experimental research designs can test causal hypotheses. Where a randomised controlled trial (RCT) includes a control arm in its design, quasi-experimental designs identify a suitable comparison group to capture what would have been the outcome had the intervention not taken place (i.e. the counterfactual), such as through propensity score matching or regression discontinuity. When data allows, and implementation of an intervention occurs at a defined point in time, an interrupted time series design could be used to evaluate the effectiveness of population-level interventions. This approach is utilised by the NIHR “BiB Breathes” project [36] which will evaluate the impact of the introduction of a Clean Air Zone on health outcomes and could be applied to assess the trend and change in selected outcomes (e.g. childhood obesity) from Connected Bradford data prior to and after the introduction of the ActEarly programme (Connected Bradford is discussed below in the section ‘data visualisations’).

###### Data visualisations

Connected Bradford links anonymous routine data across primary and secondary care with local authority social care and education datasets from the Department for Education [38]. A scoping exercise to identify the most appropriate datasets and variables is currently being carried out for the final core outcome sets for early years (COS-EY). Analysts will use secure, effective and efficient data visualisation to display actionable insights for local citizens, practitioners, commissioners and policy makers. Google Data/Looker Studio is already employed by analysts curating Connected Bradford datasets and is the most likely tool to be used for visualisations of the core outcomes as it links directly to the collected datasets. Interactive dashboards that can be filtered and sliced by demographics such as sex, ethnicity, and age at diagnosis and/or data collection date/record will be accessible.

Using timestamps on data collection, extraction or diagnosis dates and where suitable, trends over specific time periods will be displayed. Previous examples of data visualisations using related datasets include bar and line charts, data tables with filters, Radial Sets and linkage to Google Maps ‘heat maps’ by linking to lower-layer super output areas (LSOA), middle-layer super output areas (MSOA) and partial postcode information. Developers of the dashboard(s) also plan to overlay additional relevant information on dashboards, with text boxes and labels. This will be discussed with the relevant theme leads and researchers. It is possible that more definitive and direct conclusions could be afforded space in future, as well as survey or data results related to the core outcomes.

###### LifeSim

LifeSim is a computer microsimulation model that models developmental, economic, social, and health outcomes [39], and will be used to address question 3b of the meta-evaluation. The long-term benefits, and public costs of a range of different potential short-term improvements in childhood circumstances and outcomes will be extrapolated, with policy scenarios agreed in consultation with stakeholders. Two linked dynamic microsimulation models will be used: LifeSim Childhood, which models outcomes and costs from age 0 to 17 years, and LifeSim Adulthood, which models outcomes and costs from age >17 years.

Details of the LifeSim Adulthood model have been published elsewhere [39] and the development of LifeSim Childhood, using high-quality longitudinal data from the UK Millennium Cohort Study, is underway. We will undertake stakeholder consultation to agree suitable “what if” scenarios about potential improvements to a core set of childhood financial, educational and health disadvantages (what specific disadvantage measures and what potential improvement ranges). We will use administrative and survey data to describe the current demographic situation in Bradford and Tower Hamlets in terms of these childhood disadvantages. We will then use LifeSim Childhood and LifeSim Adulthood to predict the long-term benefits and cost savings of the various policy improvement scenarios.

##### Data integration

Quantitative and qualitative data will be summarised using the aforementioned methods (e.g., thematic analysis, REMs, systems mapping, data visualisations, LifeSim), and descriptive statistics will be conducted where appropriate (quantitative data). This will provide preliminary insights into the quantitative and qualitative findings independently, and inform further analyses where appropriate. Quantitative and qualitative findings will be integrated and synthesised by establishing ‘context-mechanism-outcome (CMO) configurations’ to understand what works, when, how and in what context [40]. Context-mechanisms-outcome configurations will provide a theoretical understanding of how ActEarly outcomes (O) emerge as a result of a mechanism(s) of action (M), which is/are only present in certain contexts (C) [40]. CMO configurations are informed by realist methodology, and although may be considered linear in nature, they have been used to summarise complex findings arising from whole-systems interventions [35, 41], and align with the current meta-evaluation’s overarching aim of identifying interacting contextual factors within the ActEarly’s child health system. Approaches utilised in previous research, including evaluations of whole system interventions [35, 42], will inform the methodology used in the current meta-evaluation. The current ActEarly logic model (Figure 2) outlines the hypothesised ways in which ActEarly will create change and will be used as a reference point when developing the CMO configurations. An iterative approach will be used between the qualitative and quantitative data and the ActEarly logic model to establish the CMO configurations and consider how the ActEarly logic model can be developed into an evidence-informed programme theory [42]. Such programme theory can be used to inform future work by considering the transferability of the ActEarly programme into other contexts and cities.

###### Ethical approval

The studies that form a part of the meta-evaluation have all gained ethical approval from the relevant institutional ethics boards. We have ethical approval from the UCL Research Ethics Committee for the longitudinal ActEarly Research on Research Study (2037/004) which encompasses the majority of planned meta-evaluation data collection. We are currently seeking an amendment from the UCL Research Ethics Committee to expand the scope of the longitudinal study.

### Stage 5: Measuring programme outcome and impact in terms of system changes

Stage 5 of the ENCOMPASS framework involves understanding the programme’s outcomes and impact. Following from Stage 4, the analytical strategies employed to monitor dynamic programme output will be expanded to measure programme outcomes and impact in terms of system changes. The answers and analysis for each ‘sub’ evaluation question will form a part of the wider answer to our overall evaluation questions of:

1. What major external contextual factors influenced the implementation of ActEarly?
2. How, and to what extent was ActEarly implemented as intended?
3. Is there any evidence that ActEarly has started to have a meaningful impact on early life health and well-being outcomes?

We will synthesise the information and insights gained from the analysis of the data for each of the questions, to understand the impact of ActEarly on the child health system in Bradford and Tower Hamlets.

## Discussion

This meta-evaluation protocol presents our plans of using and adapting the ENCOMPASS-framework to evaluate the system-wide impact of the early life health and well-being programme, ActEarly. The ENCOMPASS framework in its original form was focused on capturing changes in one system, while ActEarly operates across multiple systems and therefore has an additional level of complexity. We anticipate that the process of defining what is and what is not ActEarly (and what will and what will not be included in the final evaluation) will be challenging. As we are not proposing an effectiveness evaluation, we will not be able to say with certainty whether ActEarly has had a measurable impact on the child health system. However, our approach will allow us to map and explore the context, mechanisms and outcomes of implementation of a large system change intervention in order to estimate whether such implementation is able to enact systems change towards a tipping point (where ActEarly processes/intervention/collaborations continue to function independently of the ActEarly programme). It is possible that the boundaries we have defined could be contested and that imposing different system boundaries might result in different findings. Instead, it may be that our contribution will be adding to the overall understanding of how systems evaluations can (or cannot) be conducted and in identifying broader challenges in moving complex systems evaluations in public health from theory to practice.

This protocol has re-emphasised the non-linear nature of system evaluation work, and compared to a more traditional model, many of the practicalities and methodological choices that will need to be made are not as clearly laid out if compared to other designs (e.g., the evaluation protocol of a trial). This protocol aimed to describe the steps we plan to take to describe the existing system, the methods we will use and have used to define system boundaries and evaluation questions, and finally, detail some of the analytical approaches we think will be relevant in addressing those questions at this stage of the project. Due to the collaborative nature of the work, we reserve the option to change and query these choices based on feedback we receive from stakeholders to ensure that our evaluation remains relevant and fit for purpose.

For the meta-evaluation of ActEarly, the challenge will be to both assess whether there is evidence of change in the system, but also, whether we think we identified the correct levers of change. As with all research interventions (particularly of this scale), ActEarly is *part* of the system it seeks to measure and intervene in, and will without question change those systems. The difficulty comes when we want to understand and attribute changes in a system to specific interventions. This remains a key challenge of complex systems work.

### Limitations

A major challenge and limitation for the evaluation of ActEarly is the overlapping timeline between the COVID-19 pandemic and the project. We already know that the context in which the programme takes place has undergone major and unexpected changes since the project started in 2019, but we do not know if, how and to what extent the system adapted, and whether there is scope to draw apart any impact of ActEarly from the impact of the pandemic.

The baseline data for defining the ActEarly system and its boundaries was collected over several years (2018-2022) instead of a single time point. This is in part a result of the nature of the project where new themes were added to the scope until 2021, and where new projects continue to take shape today. Similarly, there have been important changes in terms of key staff and institutions involved in the research, and as such, it is difficult to define what timepoints should be defined as ‘baseline’. Boundaries and the remit of ActEarly were not clearly defined at the start of the project either, and debates regarding what is and what is not ‘ActEarly’ continue at time of writing. This could be seen as inevitable for a large public health programme that seeks to have a positive impact across a ‘whole system’, but in terms of the evaluation of the program, poses major conceptual and practical challenges that we will need to navigate going forward. So far, one of the consequences of the lack of clarity on the programme boundaries has meant that detailed and precise plans for evaluation (including the meta-evaluation questions) have had to be developed alongside the program, not in advance. We acknowledge that this could potentially introduce bias in how and what we evaluate because we already know some of the things that have/have not happened. To mediate this risk, we believe special care needs to be taken to ensure that we continue to question and reflect on our evaluation choices (incl. setting the boundaries for the evaluation) as these strongly influence what will get reported and what will not. The useful list of questions proposed by Williams [43], will act as the starting point for this process.

## Conclusion

The ActEarly meta-evaluation protocol proposes a mixed method systems evaluation of ActEarly, a large public health research programme focusing on early life health and well-being across Bradford and the Borough of Tower Hamlets (London). The recently published ENCOMPASS-framework was used as a guide to structure the proposed evaluation, with some modifications. Due to the non-linear nature of the process, we anticipate that one or more amendments to this protocol will be published in due course to describe the process through which our evaluation plans and methods are developing alongside the project itself.

## Data Availability

No datasets were generated or analysed during the current study. All relevant data from this study will be made available upon study completion.

## Notes

### Competing Interest Statement

The authors have declared no competing interest.

### Funding Statement

This work was supported by the UK Prevention Research Partnership (MR/S037527/1), which is funded by the British Heart Foundation, Cancer Research UK, Chief Scientist Office of the Scottish Government Health and Social Care Directorates, Engineering and Physical Sciences Research Council, Economic and Social Research Council, Health and Social Care Research and Development Division (Welsh Government), Medical Research Council, National Institute for Health Research, Natural Environment Research Council, Public Health Agency (Northern Ireland), The Health Foundation and Wellcome.

